# Empiric treatment with aspirin and ticagrelor is the most cost-effective strategy in patients with minor stroke or transient ischemic attack

**DOI:** 10.1101/2023.05.16.23290069

**Authors:** Kaavya Narasimhalu, Jeremy Chan, Yoong Kwei Ang, Deidre Anne De Silva, Kelvin Bryan Tan

**Author notes:** Corresponding author: Dr Kaavya Narasimhalu, Neurology (SGH Campus) National Neuroscience Institute, Outram Road, S(169608), Singapore.

## Abstract

**Background:** Patients with minor ischemic stroke or transient ischemic attacks (TIAs) are often treated with dual antiplatelet therapy regimens as part of secondary stroke prevention. Clopidogrel, an antiplatelet used in these regimens, is metabolised into its active form by the *CYP2C19* enzyme. Patients with loss of function (LOF) mutations in *CYP2C19* are at risk for poorer secondary outcomes when prescribed clopidogrel. We aimed to determine the cost effectiveness of three different treatment antiplatelet regimens in ischemic stroke populations with minor strokes or TIAs and how these treatment regimens are influenced by the LOF prevalence in the population.

**Methods:** Markov models were developed to look at the cost effectiveness of empiric treatment with aspirin and clopidogrel versus empiric treatment with aspirin and ticagrelor, versus genotype guided therapy for either 21 or 30 days. Effect ratios were obtained from the literature and incidence rates and costs were obtained from the national data published by the Singapore Ministry of Health. The primary endpoints were the incremental cost-effectiveness ratios (ICERs).

**Results:** Empiric treatment with aspirin and ticagrelor was the most cost-effective treatment regimen if the prevalence of LOF was below 7.5%. Genotype guided therapy was more cost effective than empiric aspirin and clopidogrel if the LOF was above 65.5%. Empiric ticagrelor and aspirin was cost saving when compared to genotype guided therapy. Results in models of dual antiplatelet therapy for 30 days were similar.

**Conclusion:** This study suggests that in patients with minor stroke and TIA planned for dual antiplatelet regimens, empiric ticagrelor and aspirin is the most cost-effective treatment regimen. If ticagrelor is not available, genotype guided therapy is the most cost-effective treatment regimen if the LOF prevalence in the population is more than 65.5%.

**Clinical Perspective:** *What is new?:*

- We show that empiricial ticagrelor and aspirin is the most cost-effective treatment strategy in patients with minor stroke and transient ischemic attack

*What are the clinical implications?:*

- In patients with minor stroke/ transient ischemic attack, it may not be necessary to screen for *CYP2C19* status as empirical ticagrelor and aspirin is cost effective.

## 1. Introduction

Patients with minor stroke or transient ischemic attacks (TIAs) often receive clopidogrel and aspirin in combination for 21 or 30 days as part of secondary prevention of stroke^1,2^. Clopidogrel is a platelet inhibitor that irreversibly binds to the P2Y_12_ADP receptors on platelets. Unfortunately, clopidogrel is a prodrug that needs to be metabolized into its active form by an enzymatic member of the cytochrome P450, *CYP2C19*^3^. Patients with loss of function (LOF) mutations are less able to produce the active metabolite, and therefore do not benefit from the platelet inhibition properties of clopidogrel as patients who do not have LOF mutations.

In Asians, the prevalence of LOF mutations is much higher than Caucasian patients, with up to 60% of Asians being unable to metabolize clopidogrel into its active form^4,5^. Ticagrelor, another platelet inhibitor that irreversibly binds to P2Y_12_ADP receptors, has recently been shown to be efficacious in combination with aspirin in reducing recurrent stroke in patients with minor strokes or TIA^6^. In addition, in a trial that compared ticagrelor versus clopidogrel in patients with LOF mutations, the ticagrelor group had significantly less ischemic events, with similar bleeding rates^7^.

As there is robust data now on the use of ticagrelor in patients with minor stroke and TIA, we aimed to determine the cost effectiveness of three different treatment regimens: 1. Empirical aspirin and clopidogrel, 2. Empiric Ticagrelor and Aspirin, and 3. Genotype guided therapy. We also aimed to determine how the cost-effectiveness of these treatment regimens would vary based on the LOF prevalence in the underlying population at risk.

## 2. Methods

### 2.1 Overview of the decision-analytic model

A life-long Markov model (Figure 1) was designed to simulate the outcomes of the three possible dual antiplatelet regimens. In the base model, all patients are prescribed aspirin and clopidogrel for 21 days, followed by lifelong aspirin. In Model A, all patients are prescribed aspirin and ticagrelor for 21 days, followed by lifelong aspirin. In Model B, all patients undergo *CYP2C19* testing, and those who are LOF carriers are prescribed aspirin and ticagrelor for 21 days then lifelong aspirin, while those with no LOF mutations are prescribed aspirin and clopidogrel for 21 days then lifelong aspirin. Clopidogrel was given in a loading dose of 300 mg followed by a 75 mg dose daily. Ticagrelor was given in a loading dose of 180 mg followed by a 90 mg dose twice a day. Aspirin was given in a loading dose of 300mg, followed by a dose of 100mg once a day. Analyses models were also rerun with dual antiplatelets for 30 days instead of 21 days based on the THALES trial^6^.

**Figure 1.**
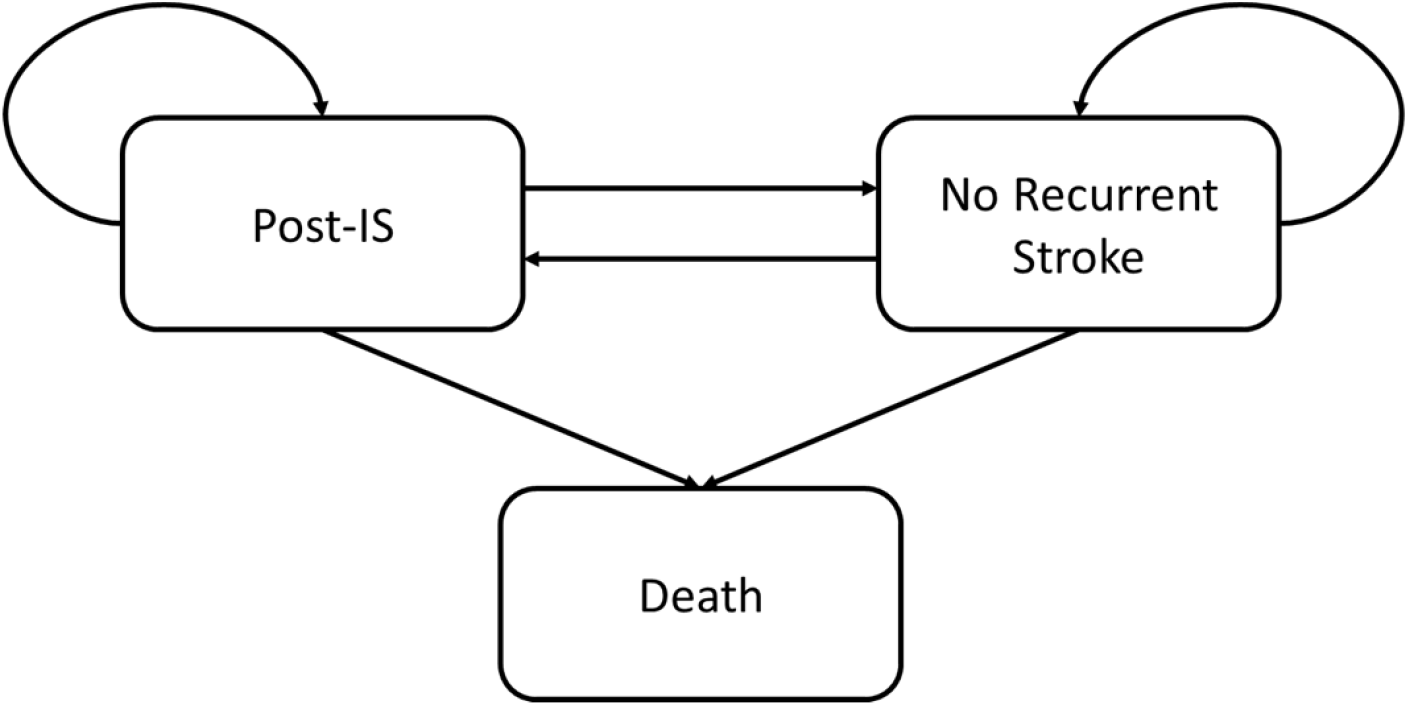
Schematic diagram of Markov model used in the analysis.

In the genotype guided therapy arm, patients were categorized by *CYP2C19* genotype as carriers or non-carriers of LOF alleles. LOF alleles included *CYP2C19*2, CYP2C19*3, CYP2C19*4, CYP2C19*5, CYP2C19*6, CYP2C19*7, CYP2C19*8, CYP2C19*9* and *CYP2C19*10. CYP2C19*1* and *CYP2C19*17* alleles were the wild-type allele and gain-of-function allele respectively^4^. Carriers are defined as patients who carry at least one of the LOF alleles while non-carriers do not carry any of the LOF alleles.

### 2.2 Model cohort

The population modelled was a hypothetical cohort of 65-year-old Singaporean patients who suffered their first ischemic stroke. The model was ran for yearly cycles until age 85 to observe the lifetime costs and quality-adjusted life years (QALYs) of each treatment strategy^8^.

### 2.3 Model parameters

All model inputs were derived from published literature or locally-sourced data from the Ministry of Health, Singapore and hospitals. The parameters are summarized in Table 1. We used the local background rates of ischemic stroke and mortality at every age and multiplied them by the risk ratios and standardized mortality ratio to derive the transitional probabilities in the Markov model. The background rates and risk ratios were obtained from published government reports^9,10^ and past studies on stroke patients respectively^7,11–17^. Specifically, the government reports include the Singapore Stroke Registry^9^ that was compiled by Health Promotion Board, Singapore who collected epidemiological and clinical data on stroke cases in Singapore public hospitals between 2007 and 2019. Additionally, the background mortality rates at every age were obtained from the Complete Life Tables^10^ for Singapore Resident Population 2017-2018 that was compiled by the Department of Statistics, Ministry of Trade and Industry, Singapore.

**Table 1.**
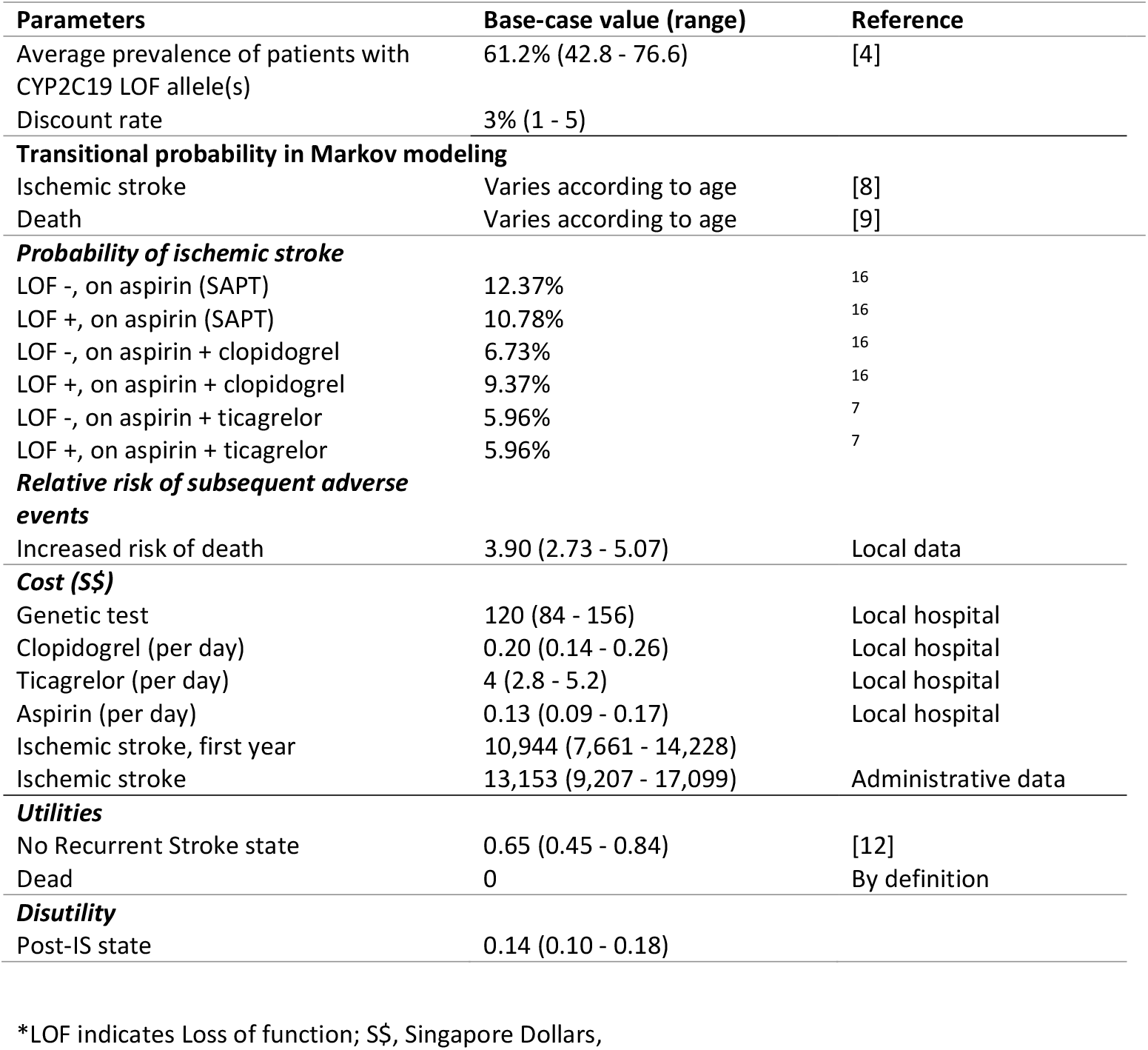
Model input used in comparison of antiplatelet strategies after incident stroke.

The prevalence of LOF allele carriers was previously reported in 506 genomic samples of healthy Singaporeans from the major local ethnic groups^4^. We obtained the probabilities of developing a recurrent ischemic stroke among carriers and non-carriers taking clopidogrel from a Chinese study that evaluated the outcomes of 625 patients^11^. The risk ratios were then calculated by dividing the reported numbers with the probabilities of a healthy Singaporean developing stroke at the reported average age in the study. A similar method was applied to a subgroup analysis of the CHANCE 2 trial used to obtain the risk ratios of patients developing a recurrent ischemic stroke while taking ticagrelor^7^. We assumed that the pharmacological effects of ticagrelor were not dependent on the patients’ genotype^18^. As these trials reported rates of outcomes until the 90 day follow up, all patients were assumed to have the same risk of stroke from day 90 to day 365 while on aspirin monotherapy based on the difference between the 1 year follow up data^17^ and the 90 day follow up data^16^ from the CHANCE study. The proportion of patients who develop disabilities after suffering from a stroke and the standardized mortality ratio used in the study was obtained from the Ministry of Health, Singapore.

### 2.4 Utility and cost inputs

The utilities used for this study were measured in QALYs to reflect the state of one’s health. Patients who develop a stroke can be disabled or not disabled. Due to the lack of locally derived data on the utility of patients after suffering from a stroke, we followed the utility scores outlined in another cost-effectiveness study on stroke patients^19^.

All costs were estimated in Singapore dollars (S$). We accounted for inpatient, outpatient, and pharmaceutical costs using a local healthcare provider’s perspective. The cost of the *CYP2C19* genotyping test was S$120 and was assumed to be applied once only. Additionally, the sensitivity and specificity of the test were assumed to be 100%^20,21^. The cost of clopidogrel is S$0.20 per day and the cost of ticagrelor is S$4 per day. Ischemic stroke costs were obtained from administrative data from the Ministry of Health, Singapore.

### 2.5 Cost-effectiveness analysis and sensitivity analysis

Cost-effectiveness and sensitivity analyses were undertaken using Microsoft Excel for Office 365 (Microsoft, Redmond, Washington, USA). The Incremental Cost Effectiveness Ratios (ICER) were calculated dividing the differences in costs between the base model and the two alternative strategies, over the differences in Quality Adjusted Life Years.

As there is no official willingness-to-pay threshold for adopting health technologies in Singapore, we chose to implement a threshold of the gross domestic product per capita as recommended by World Health Organization is commonly used in publications^22^, resulting in a threshold of willingness to pay of S$60,000/QALY (based on 2020 data).

Sensitivity analyses were conducted to evaluate the robustness of the results from the cost-effectiveness model. One-way sensitivity analysis was performed over the parameter ranges. The distribution and parameter ranges were derived from the data reported in the sources for these parameters, with lognormal, beta and gamma distributions used where source data is available as recommended by the CEA literature. For costs data, where distribution is not reported, we use a triangular distribution of ±30% to identify parameter inputs that may impact the results. To assess the uncertainty of all input parameters in combination, a probabilistic sensitivity analysis was performed with 10,000 Monte Carlo simulations by using random values between the upper and lower range of each model input. We also modeled the costs, QALYs, and ICERs based on hypothetical populations with a range of LOF prevalence from 0 to 1.

## 3. Results

### 3.1 Base case analysis

In a population of patients with minor ischemic stroke or TIA presenting at age 65 years, model A in which patients are prescribed empiric aspirin and ticagrelor was the most cost-effective model, being both cheaper and with a higher QALY (Table 2). This was irrespective of whether dual antiplatelet therapy (DAPT) was for 21 or 30 days. Empiric aspirin and clopidogrel was more cost effective than genotype guided therapy at the LOF prevalence of 61.2% in Singapore.

**Table 2.** Expected costs, quality-adjusted life years and incremental cost-effectiveness for model A (aspirin+ clopidogrel), model A (aspirin+ ticagrelor) and model B (genotype guided therapy) in models with dual antiplatelet therapy for 21 and 30 days respectively.

| Strategy | Cost (S\$) | QALYs | Incremental | | | Number of Ischemic Strokes Prevented |
| --- | --- | --- | --- | --- | --- | --- |
| | | | Cost (S\$) | QALYs | Cost/QALY (S\$) | |
| **<u>DAPT for 21 days</u>** |  |  |  |  |  |  |
| Base Model | 26,098 | 5.1229 | - | - | - | 1.481 |
| Model B | 26,232 | 5.1250 | - | - | - | 1.479 |
| Model A | 26,138 | 5.1253 | - | - | - | 1.478 |
| A vs Base | - | - | 40 | 0.0024 | 16,395 | - |
| B vs Base | - | - | 134 | 0.0021 | 63,426 | - |
| B vs A | - | - | Dominated by A |  |  | - |
| **<u>DAPT for 30days</u>** |  |  |  |  |  |  |
| Base Model | 26,100 | 5.1229 | - | - | - | 1.481 |
| Model B | 26,254 | 5.1250 | - | - | - | 1.479 |
| Model A | 26,174 | 5.1253 | - | - | - | 1.478 |
| A vs Base | - | - | 74 | 0.0024 | 30,590 | - |
| B vs Base | - | - | 155 | 0.0021 | 73,362 | - |
| B vs A | - | - | Dominated by A |  |  | - |

\*S\$ indicates Singapore Dollars; QALYs, Quality-adjusted life years; DAPT, dual antiplatelet therapy.

### 3.2 Sensitivity analyses

The results of one-way sensitivity analyses are summarized in Supplementary Table 1. Supplementary Figure 1 presents the results in a tornado diagram in decreasing order of how sensitive the ICER is to each parameter with Supplementary Figure 1A summarizing the sensitivities when comparing model B to model A, and with Supplementary Figure 1B summarizing the sensitivities when comparing the model B to the base model.

### 3.3 Probabilistic sensitivity analyses

Results from the probabilistic sensitivity analyses suggest that both incremental costs and incremental QALYs fluctuate substantially when we varied the parameters in combination but remain cost effective in all scenarios (Supplementary Figure 2 A & B).

### 3.4 Variation of model function across LOF frequency

Figure 2 summarizes the costs, QALYs, and ICERs of the three different treatment strategies across different prevalences of LOF for models that use 21 days of DAPT (Figure 2A) and 30 days of DAPT (Figure 2B). Results from these analyses show that empiric ticagrelor and aspirin (model A) is the most cost-effective strategy when the LOF prevalence is more than 7.5% for models considering DAPT for 21 days, and 24.2% for models considering DAPT for 30 days. When comparing empiric aspirin and clopidogrel (base model) to genotype guided therapy (model B), empiric treatment with aspirin and clopidogrel was more cost effective in populations where the LOF prevalence is below 65.5% in models of DAPT for 21 days and in populations where the LOF prevalence is below 83.9% in models of DAPT for 30 days. Empiric ticagrelor and aspirin is cost saving when compared to genotype guided therapy.

**Figure 2:**
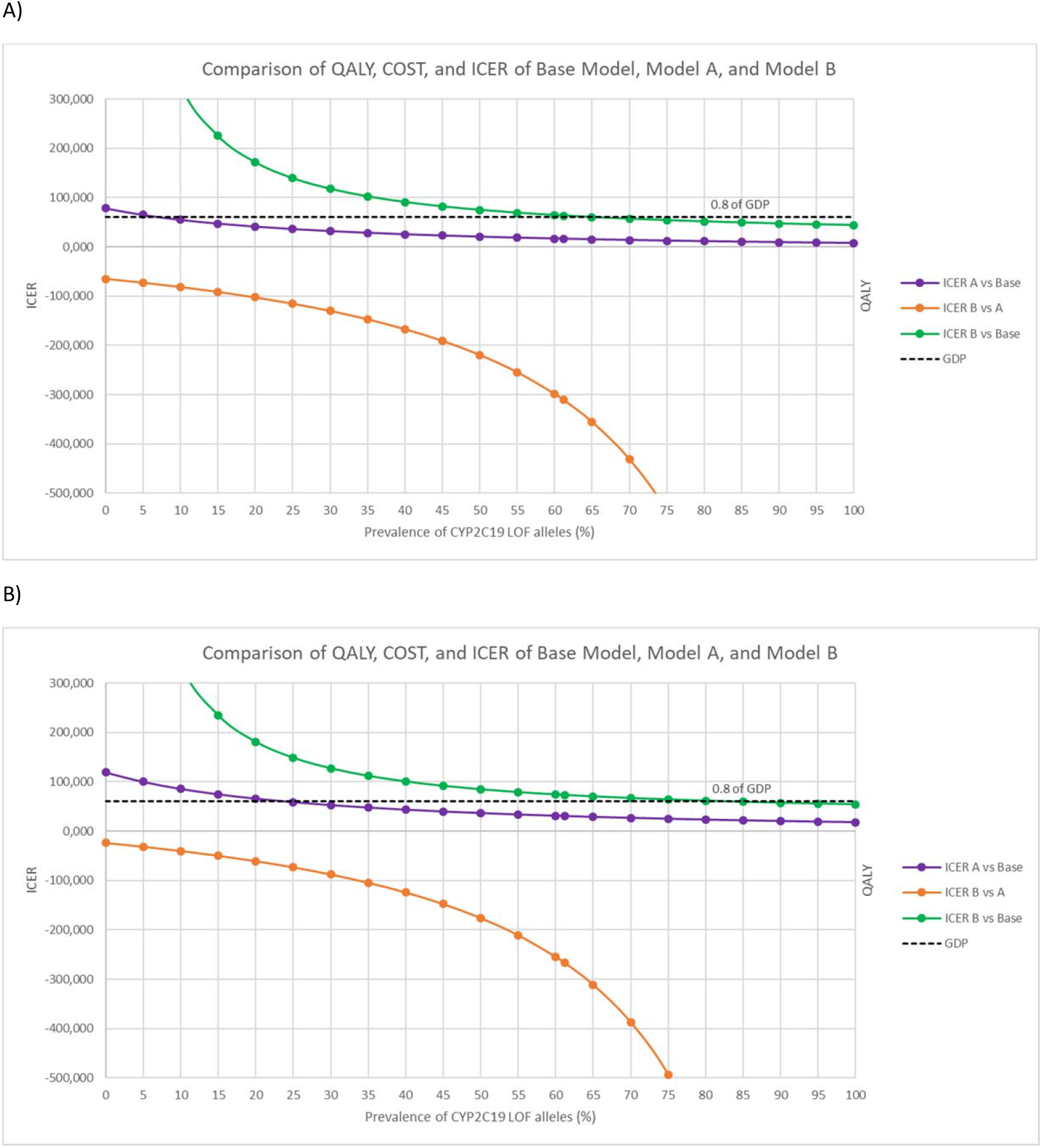
Variance of costs, quality of life years (QALY), and incremental cost effectiveness ratios (ICERs) of the different treatment models across the prevalence of loss of function alleles for 21 days (A) and 30 days (B).

## 4. Discussion

In this study, we demonstrated that empiric ticagrelor and aspirin is the most cost-effective treatment regimen in patients with minor stroke and TIA in most populations. We were also able to show that in situations where ticagrelor may not be available, for models were DAPT is for 21 days, empiric aspirin and clopidogrel is cost effective in populations with LOF prevalence less than 65.5%.

A previous study from our group^23^ showed that it was cost effective to screen for LOF mutations in ischemic stroke populations who are planned for single antiplatelet therapy, with ticagrelor as a substitute for clopidogrel in those with LOF mutations. However, it did not focus on patients with minor stroke and TIA in whom dual antiplatelet therapy is indicated. The shorter duration of DAPT (21 or 30 days) may explain why empiric ticagrelor and aspirin rather than genotype guided therapy is more cost effective in the present study. One previous group from China^24^ has shown that genotype guided dual antiplatelet therapy is more cost effective than standard medical therapy by risk stratifying by clinical functional status after the first stroke. A Canadian group^25^ has previously shown that genotype guided therapy with patients with LOF mutations being given ticagrelor is cost effective at a threshold of

$50,000 CAD per QALY. Yet another study has previously demonstrated that the addition of cilostazol to aspirin or clopidogrel is cost effective in patients with non-cardioembolic strokes^26^. However, no previous stroke studies have compared cost effectiveness of genotype guided therapy with empiric ticagrelor and aspirin.

The THALES^6^ trial showed a significant reduction in the composite endpoint of stroke or death in patients treated with aspirin and ticagrelor compared to aspirin alone in patients with mild to moderate ischemic strokes. A pre-specified substudy of the THALES trial^27^ has also shown that patients with ipsilateral atherosclerotic disease of at least 30% benefited from the addition of ticagrelor to aspirin. As such, despite a slightly increased risk of bleeding^6^ when compared to clopidogrel, ticagrelor now has been discussed as one of the antithrombotic agents used for secondary stroke prevention in the 2021 AHA guidelines^28^. More recently, the CHANCE-2 trial^7^ has shown that ticagrelor is an acceptable alternative to clopidogrel in combination with aspirin in secondary stroke prevention for patients with minor stroke or TIA who carry LOF alleles.

There were some limitations in this study. Firstly, we modelled only direct costs and no accommodation was made for indirect costs (adverse drug reaction, transport, time, cost of work lost). Secondly, our model was developed from a Singaporean healthcare perspective, and as such, the costs and probabilities may differ from other countries/regions and therefore affect the generalizability of these results. Thirdly, our model incorporates event rates from mainly Chinese studies (CHANCE and CHANCE 2) and as such, the results may not be generalizable to non-Chinese populations.

## 5. Conclusion

This study suggests that in patients with minor strokes or TIAs, empiric ticagrelor and aspirin therapy is the most cost-effective. If ticagrelor is not available, then genotype guided antiplatelet therapy is cost-effective if the populations LOF prevalence is more than 65.5% for 21 days of DAPT and 83.9% for 30 days of DAPT.

## Data Availability

Data used to general these models are from publically available trial data. All model inputs are listed in Table 1.

## Non-standard Abbreviations and Acronyms

TIA: transient ischemic attack
LOF: loss of function
ICER: incremental cost-effectiveness ratio
QALY: quality-adjusted life years
S$: Singapore dollars
DAPT: dual antiplatelet therapy

## Acknowledgements

None

## Author contributions

Study concept and design KN, KBT. Acquisition of data KN, JC, YKA. Analysis and interpretation of data KN, JC, YKA, KBT. Drafting of manuscript KN. Critical revision of manuscript for important content -all authors. This study was unfunded. KN and JC had access to all data and take ownership for the integrity of the data and analyses.

